# Evaluation of High Flow Local Extraction on control of the aerosol plume in an operating theatre

**DOI:** 10.1101/2021.05.16.21257155

**Authors:** Logan Marriott, Matthew Harper, Tongming Zhou, Chenlin Sun

**Affiliations:** Consultant Anaesthetist, Fiona Stanley Hospital, Western Australia, Australia; Professor, Faculty of Engineering and Mathematical Sciences, University of Western Australia, Australia; PhD Student, Faculty of Engineering and Mathematical Sciences, University of Western Australia, Australia

**Keywords:** aerosol, high flow local extraction, aerosol-generating procedure, tracheal intubation, SARS-CoV-2, COVID-19, plume, personal protective equipment, engineering controls

## Abstract

**Background:** Engineering controls are a necessity for minimising aerosol transmission of SARS-CoV-2, yet so far, little attention has been given to such interventions. High flow local extraction (HFLE) is a standard in other industries that deal with airborne contaminants.

**Objective:** This study aims to provide a quantitative evaluation of an HFLE concept feasible to implement in most real clinical settings.

**Design:** A unique combined experimental model of Laser sheet illumination videography paired with continuous nanoparticle counts was used to quantitatively assess the impact of HFLE in an operating theatre. Propylene Glycol was aerosolised via a customised physiological lung simulator and dispersion was measured in 3 dimensions. Cumulative probability heat maps were generated to describe aerosol behaviour. Continuous particle counts were made at 15 locations throughout the room to validate laser assessments.

**Results:** High flow local extraction reduced dispersion of simulated exhaled aerosols to undetectable levels. With the HFLE in operation and optimally positioned, the aerosol plume was tightly controlled. Particle counts remained at baseline when HFLE was active. HFLE becomes less effective when positioned at increasing distance from the mouth.

Aerosol plume behaviour in the absence of HFLE was highly variable and unpredictable.

**Conclusions:** This analysis demonstrates great potential for HFLE to have a significant impact in reducing aerosol transmission. Simple HFLE devices can be easily engineered and could be widely deployed without impacting on the safe delivery of care.

## Introduction

There is increasing evidence that aerosol transmission plays a significant role in the spread of the SARS-CoV-2 (1,2). Aerosols are variably defined by size but are generally accepted to be particles of less than 5μm. They are distinct from larger droplets which tend to settle on surfaces within seconds to minutes, traveling significant distances suspended in air near indefinitely (3) and can remain viable for prolonged periods (4). Health care workers are overrepresented in the infected population, and have at least a 3 fold increased risk of becoming infected in comparison with the general population (5). Repeated and prolonged exposure to infected individuals correlates positively with severity of illness in health care workers (6).

Aerosol generating procedures (AGPs) in medicine may present an increased risk of transmission to health care workers despite effective PPE discipline (7,8). However, it has been suggested that simply breathing and talking represent significant aerosol transmission risks (9). Indoors, aerosol based “viral load” in clinical areas is determined by content released; room size for dilution; and airflow clearance for washout (10). Recent recommendations for adaptations to ICU practice reflect this understanding and include configurations that allow the nursing staff to monitor the patient from outside the room (11), (12).

The Centers for Disease Control and Prevention (CDC) places engineering controls of risk (such as optimized ventilation) above PPE in its hierarchy of controls (Supplementary Figure 1). Despite this, high flow local extraction of exhaled aerosols from close to the patient’s airway is a concept that has received little attention so far as a strategy to reduce transmission risk to health care workers (13), (14). Measuring the efficacy of such devices is challenging and may be a contributing factor in this. Furthermore there is no currently available commercial device validated for this purpose in the clinical setting.

In this article we describe a simple, prototype high flow local extraction device and an experimental model developed to determine its efficacy in an operational clinical environment. The experimental protocol is refined from the work of previous investigators.

Previous assessments of aerosol behaviour have used a high powered laser light sheet to illuminate a smoke plume (10,15–22), via a camera at right angles or schlieren imaging (23). Other investigators have used particle counters to quantify a single location measurement of an aerosol surrogate such as nebulised saline and powders in low fidelity simulation (24,25). The protocol described here uses aerosolised propylene glycol as a surrogate for respiratory aerosols introduced by a physiological lung simulator into a normal operating theatre (26–28). Assessments of aerosol behaviour and spread are made using a combination of laser sheet illumination and nanoparticle meter counting at multiple locations in the room.

## Methods

### Experimental model

#### Physiological Lung Simulator

A physiological negative pressure lung simulator (Referred to herein as “test apparatus”-see Appendix 2), was developed specifically for this project in order to create physiologically realistic and repeatable breathing patterns. Airflow from the test apparatus was passed directly into the distal airway of a TrueCorps mannequin head^(a)^ with anatomically accurate adult human oro and nasopharyngeal anatomy, to simulate realistic plume airflow. Flow volume loops were measured using a Hamilton T1 ventilator flow sensor^(b)^. Tests were conducted in an operating theatre, under laminar flow (meeting healthcare department performance requirements), with all normal theatre equipment present.

#### Aerosol Generation

Propylene glycol was chosen as a simulated respiratory aerosol. Particle sizes of propylene glycol measure in the range of 145-175nm once vapourized, being slightly greater than a single SARS-Cov-19 virus, and are neutrally buoyant, hence will move in airflows in a way representative of particles generated by aerosol generating procedures.

#### HFLE Prototype

The prototype high flow local extraction device was designed to redirect existing building extraction and filtration systems to an intake close to the patient. The 3 principal components are as follows:

1. Building ventilation intake shroud (an adaptable structure that can affix to a room’s ventilation extraction duct opening). In this case a stainless steel box was prototyped to match the size of the local building extraction outlet duct. It was affixed to the surrounding wall using standard duct tape.
2. Flexible ducting, in this prototype conventional 15cm tumble dryer hosing was used.
3. Patient-end intake cowling (an ergonomically shaped capture funnel, prototyped in cardboard, that sits close to the patient’s face without obstructing airway access) to improve airflow entrainment.

HEPA filtration is integrated into the building’s ventilation system immediately downstream of the intake shroud. There are no moving parts required and the device tested here is easy to construct from simple materials that are widely available.

(See Appendix 3, figures 8-10)

#### Test Setting

Tests were conducted in an operating theatre with normal positive pressure laminar flow, of volume 177m3 (8×7.4×3), and 39 air changes per hour (ACH) delivered through eight central ceiling air HEPA diffusers. Air was extracted through four floor level vents each equipped with HEPA filters. Flow through the prototype high flow local extraction device was measured to be 62L/sec corresponding to a velocity of 3.5m/sec at the intake cowling. The test apparatus was placed in the standard operating room position for intubation on an operating table, 110cm from the floor, centered under the laminar flow. Normal laser safety precautions were in place and additional blackout shielding added to prevent ambient light artefact and contaminant laser scatter.

#### Operational Parameters

Ventilation patterns of 1200ml tidal volume (Vt), and respiratory rate (RR) 12 were used to simulate maximal tidal breathing, as is common prior to the induction of anaesthesia. Furthermore, this pattern represents a greater challenge to the performance of the HFLE device than standard tidal breathing. Propylene glycol was introduced as previously described, to act as an aerosol tracer. A single test consisted of a minimum of 5 respiratory cycles as outlined above, with a constant supply of power to the heating coil of the aerosol generator throughout.

#### Laser Illumination Planes

Throughout each test, a 5-Watt Argon-ion laser was directed across the propylene glycol aerosol plume to map its characteristics and behaviour. A high speed video camera^(f)^ with a pixel resolution of 1920 x 1080 and a frame rate of 50fps was employed to record video of the aerosol plume during tidal breathing, filming perpendicular to the sheet of laser light.

Tests were completed in 11 axial slices and 9 sagittal slices, at 5cm increments relative to the centre of the mannequin’s mouth (see Figure 1).

**Figure 1:**
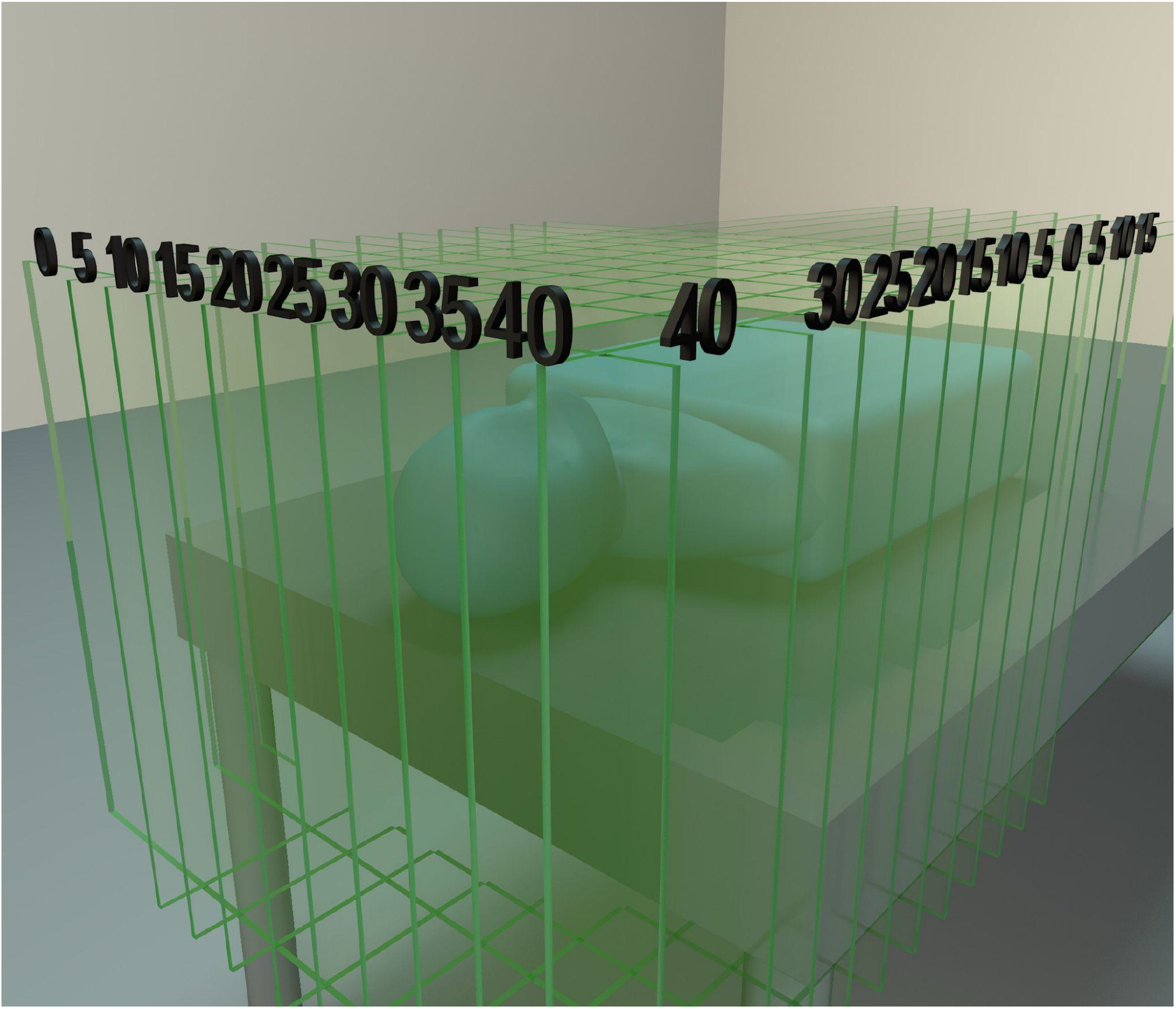
Schematic of Laser assessment grid, showing sagittal and axial slices used. Link text : Tests were completed in 11 axis

This whole sequence was repeated with the HFLE intake cowling repositioned at 5cm increments from the mannequin’s lips in a caudal direction from 15cm to 35cm.

#### Video Image Analysis

The video frames were converted to grey-scale using proprietary coding for probability analysis, where pixel intensity signified particle concentration. A reference image was taken before each test. Subtracting the reference image gave a resulting probability plot of aerosol dispersion. Five breaths of cumulative data were used to produce a heat map.

#### Particle Count Testing

Particle count testing was conducted in parallel with laser imaging, to quantify the variation of the aerosol in the room with time and to assess the impact of high flow local extraction on plume control. Particle concentrations were measured continuously using a TESTO DiSCmini particle counter^(g)^ (particles 10-700nm absolute range see Appendix 2). Data was sampled at a rate of one per second and was digitally logged throughout each test. Plots were made using Matlab. The intake cowling for high flow local extraction device was positioned optimally for extraction, 20cm from the mannequin’s mouth, as dictated by the laser imaging experiments (see results). Particle measurements were made continuously throughout each test, which consisted of five distinct phases, detailed in Supplementary Table 1. As shown, an initial five minute phase of background particle measurement preceded any artificial aerosol generation, to serve as a control, confirming values were less than 500 particles/cm^3^. During each aerosol generation phase, 4ml of propylene glycol was aerosolized completely through the test apparatus.

Particle tests were conducted at 15 different locations relative to the mannequin head (Supplementary Illustration 1).

## Results

### Laser Image Analysis

Without high flow local extraction, the plume can be observed to travel in a cephalad direction under the influence of laminar flow, (Appendix: Supplementary Figure 2). The images, video and heat maps demonstrate that under these conditions the plume is not stable and varies in both shape and distribution from breath to breath.

By reversing the position of the patient, it can be seen that the laminar flow directs the plume caudally. This simple intervention may help reduce risk to the health care workers standing by the mannequins head. (Figure 2).

**Figure 2:**
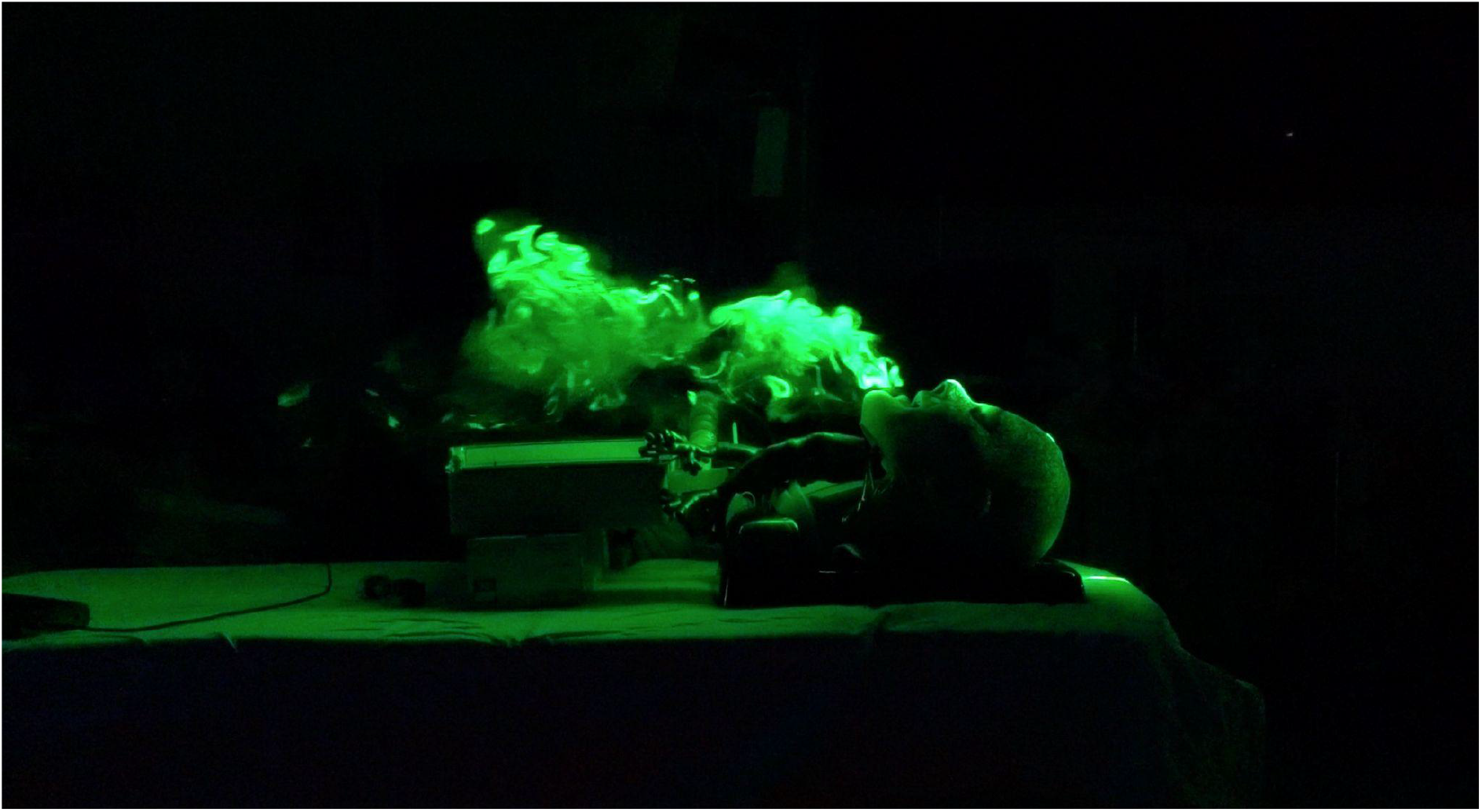
Patient position re-oriented to be located at the edge of the laminar flow, allowing the proceduralist for AGP to stand directly under the clean upstream flow. Laser image pre-data processing, at midline in the Sagittal plane, showing plume dispersing caudad, away from the HCW. Link text : This simple intervention may

The probability heat-maps shown in Figure 3A further characterise the behaviour of the plume in sagittal and axial planes. The heatmaps in Figure 3B demonstrate the effect of high flow local extraction in operation positioned at 25cm from the mouth. FIgure 3C shows a single slice at 15cm offset in the sagittal plane, comparing HFLE (left), to standard airflow (right). The plume can be seen dispersing throughout the whole grid in a cephalad direction and was noted to vary from breath to breath in the latter.

**Figure 3:**
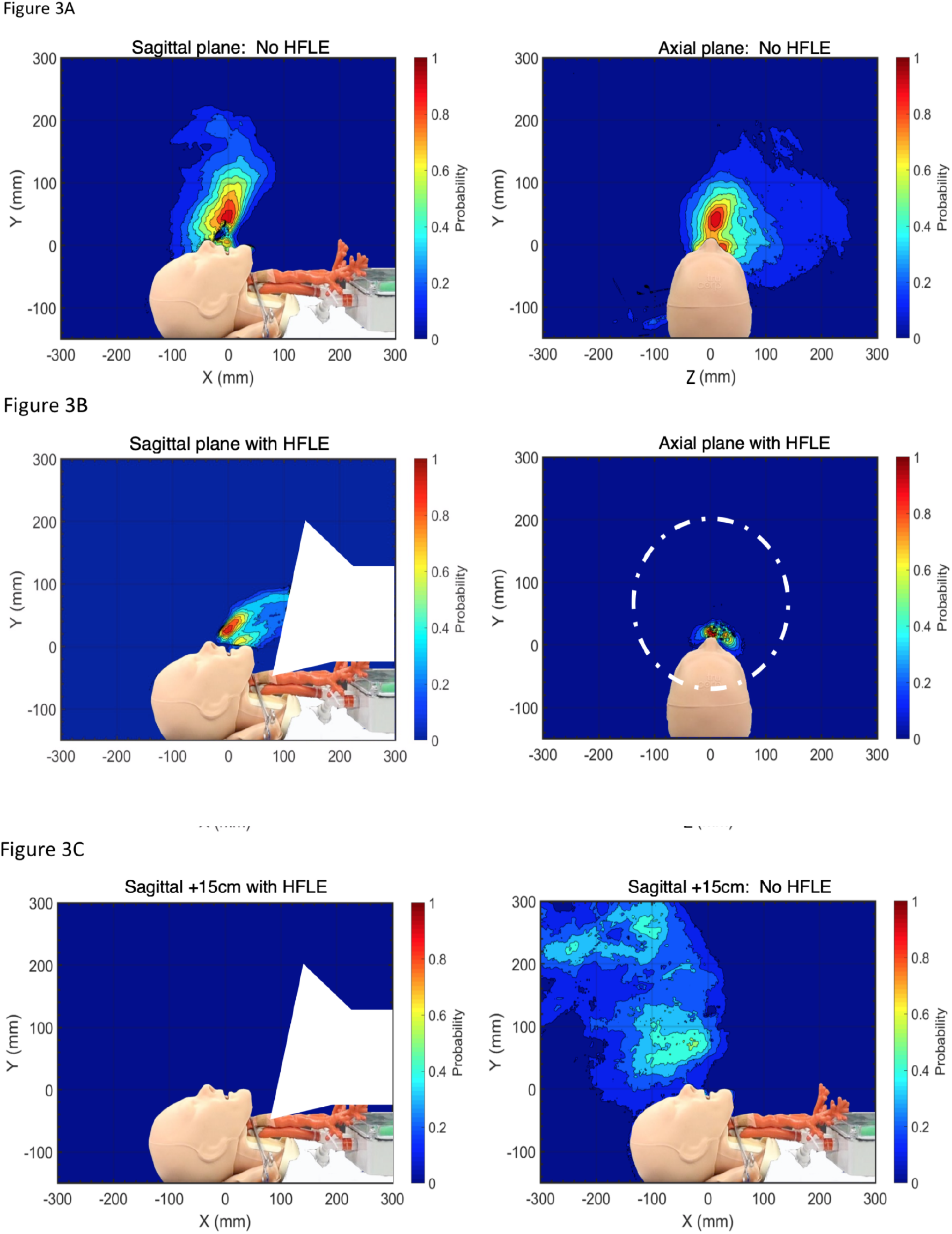
Heat maps accumulated through 5 respiratory cycles in sagittal and axial planes. (a) midline, under standard airflow conditions. (b) midline, with HFLE device positioned 25cm caudad from the mouth, (c) sagittal views 15cm from midline comparing HFLE (left), to standard airflow (right) Link text : further characterise the behaviour

Without high flow local extraction the probability of encountering aerosol in the studied area is variable, but significant, ranging from 0-100% depending on position. No safe location could reliably be identified in the absence of HFLE (Figure 4).

**Figure 4:**
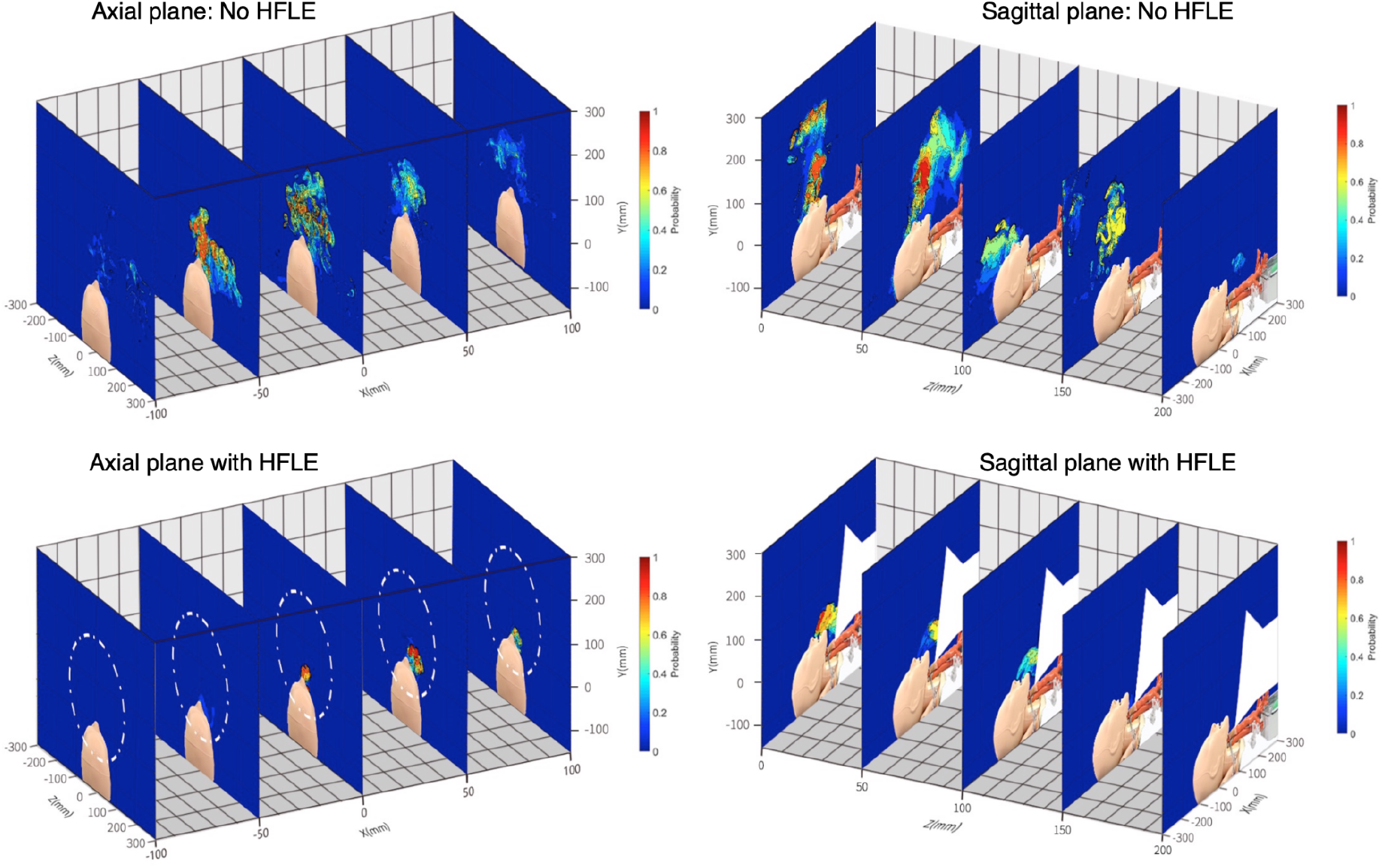
Probability plots of aerosol plume in both Axial and Sagittal planes at 5cm increments. Without HFLE (top). With HFLE - device indicated in white (bottom) Link text : No safe location could

With the high flow local extraction device positioned within 25cm of the vapour origin, the plume is entirely controlled and extracted with no measurable aerosol escape. The probability of encountering aerosol outside of the observable extraction jet is 0%. Efficacy is inversely related to distance from the source. It can be observed from video in the axial plane, when the high flow local extraction device is positioned at 35cm caudally from the mouth, significant aerosol escapes laterally away from the extractor intake and disperses into the room.

### Particle meter

Significant peaks in measured particle counts were observed in several locations within the operating theatre. These peaks correlated closely in time with phases of aerosol generation.

The highest observed increase in particle concentration was seen in the sagittal axis at 170cm from the mouth and 160cm above the floor (Figure 5: Particle Count at Location A). Particle counts reached over 30000/cm^3^ (over 300x baseline).

**Figure 5:**
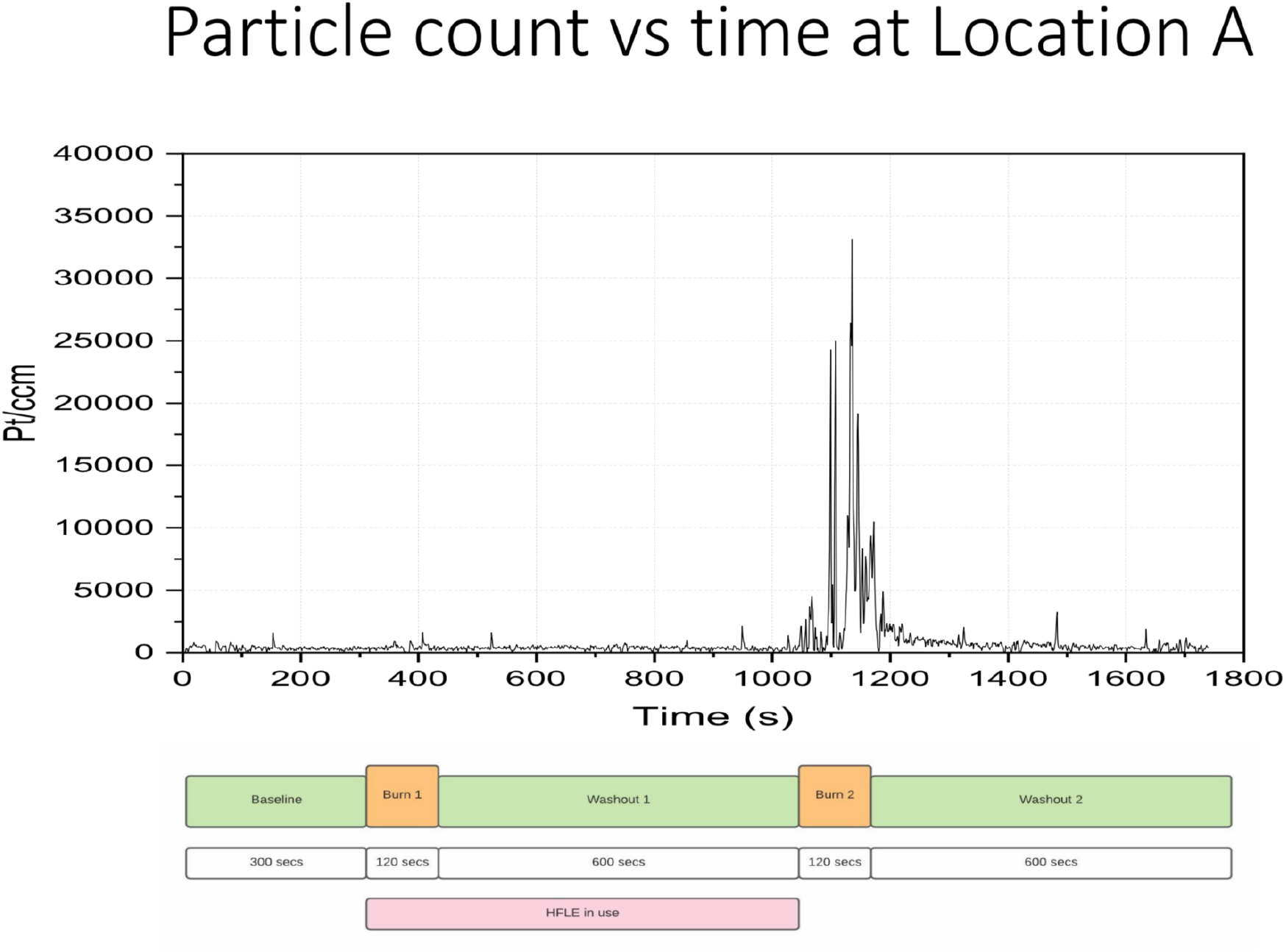
Continuous particle meter count throughout continuous test sequence at location A; Sagittal plane, 170cm distance, 160cm height. HFLE positioned 20cm caudad from mouth Link text : The highest observed increase

Particle counts throughout much of the rest of the room were highly variable and dependent upon location within the airflow. Indeed, some locations did not record any measurable particles above baseline, presumably due to air currents bypassing this location.

During the use of high flow local extraction (positioned at 20cm from the mouth), particle counts were undetectable above baseline values at all room locations tested. These findings strongly support the quantitative assessments made by laser.

## Discussion

This article has demonstrated that the prototype high flow local extraction device as tested, is highly effective in this clinical setting, reducing dispersal of exhaled aerosols to undetectable levels. Furthermore, the technique described, provides the first in-situ, scientific assessment of the efficacy of high flow local extraction in simulated aerosol capture.

Utilising the physiological negative pressure lung simulator (test apparatus) we have demonstrated a robust experimental model that is able to deliver repeatable, measurable physiological breathing patterns. The laser measurement techniques used here build on the contributions of previous work to more fully characterise quantitatively and qualitatively the behaviour of an exhaled plume in a clinical environment. In parallel, continuous particle measurements made during controlled aerosol release, support and complement these findings (13,16,24). Whilst the static images presented and statistics of particle counts are useful for quantitative analysis, a much richer painting of aerosol behaviour can be achieved by viewing the dynamic video of these experiments.

We have also described extensively the behavior of the plume under laminar flow, and have observed simple mitigation strategies that could be implemented immediately to reduce the risk of aerosol exposure to health care workers. One key principle of Centers for Disease Control and Prevention guidance is for health care workers to stand upstream in the airflow from the patient source to reduce risk of exposure. Our observations of plume behaviour in laminar flow support this advice. A simple mitigation measure in the operating theatre would be to reverse the position of the table during aerosol generating procedures, with the patient’s feet pointing out from the centre of the room.

The CDC/NIOSHs hierarchy of controls states that controlling exposure to occupational hazards is the fundamental method of protecting workers. Much of the global discussion on risk mitigation has focused on PPE, yet this guidance explicitly states that engineering controls should be favoured above PPE because they are designed to isolate the hazard (Supplementary Figure 1).

However PPE, even when used correctly, represents the last line of defence (29) and is far from infallible. Masks at best, are not 100% effective and there are shortcomings in both individual fit and shortages in supply (30).

Engineering controls such as negative pressure rooms isolate the risk to surrounding areas, but these rooms offer no additional protection to the staff within, beyond their air cycling capability, and should be considered “hot” zones. Further attempts to create engineering controls that limit exposure have led to a proliferation of medical innovations targeted towards reducing exposure of health care workers especially during AGPs eg aerosol box etc. Such innovations have generated much interest, but have rightly been criticised in the peer reviewed literature for failing to satisfy the requirement for validative assessments of efficacy (31), (32), (24), (33), (34). In particular, containment boxes do not remove the risk, but just store, concentrate and delay aerosol release into the room airspace. The addition of suction evacuation however, has shown measurable reduction in moderate sized particles outside of a tested hood in low fidelity (25).

Current literature on aerosol behaviour in the clinical settings is largely underdeveloped, with only several key investigations being most cited (15,21), and current SARS-CoV-2 related studies to date are of low fidelity. Common weaknesses include inadequate replication of physiological breathing/coughing and the substitution of nebulised liquid droplets for aerosols, and measurement in a single location often upstream of the plume.

The importance of further investigations of aerosol behaviours in hospitals cannot be overstated. SARS-CoV-2 has been detected well beyond the distance of droplet spread throughout the hospital, including areas without direct clinical exposure. (35,36) Poor understanding and implementation of building ventilation systems can potentially exacerbate spread. There are clear guidelines outlining the key safety principles of building ventilation for aerosol precautions (37–39). The CDC & WHO recognise that the first step for environmental risk mitigation in tuberculosis patients is local exhaust ventilation, and to avoid airflow designs that cause “short-circuiting” (Appendix 1, Supplementary Figure 3).

There are devices (see appendix 3) purporting to scavenge particles at much lower flow rates than HFLE, but as yet they remain unvalidated and provide no efficacy data to support their routine use at this time. The only previous assessment of high flow extraction in the medical literature demonstrated an effective reduction in simulated aerosol levels (13). This, inline with the CDC/NIOSH recommendations has been largely overlooked in major guidelines on the management of airways in patients with COVID-19 (14,40).

Indeed, medicine is starkly at odds with other industries where workers are at risk from airborne contaminants. The construction and welding industries have enshrined “Local Exhaust Ventilation” in workplace safety guidance as an engineered control for decades (safeworkaustralia.gov.au, breathefreely.org.uk) Medical colleges contribute to such guidance yet seem unable to apply the same standard for healthcare workers. It is certainly not inconceivable that the overrepresentation of healthcare workers in the infected population, is at least in part attributable to poor building ventilation/extraction and chronic exposure. The long term impact of this occupational hazard will be hard to quantify.

Our study, whilst we believe it to be a sound proof of concept, undeniably has some limitations. The use of propylene glycol in lung simulator apparatus as a surrogate for true respiratory aerosols produced in vivo, must be acknowledged. However, this approach was chosen to enable the control of variability in respiratory dynamics observed in human volunteers. Video of human volunteers exhaling propylene glycol and using the HFLE device (without quantitative assessment) is available (https://youtu.be/XA6EGgDhX24) and demonstrates similar findings. Our experiments were limited to the study of aerosols in the range of 140-175nm. The behaviour of larger particles (large respiratory droplets) in this context may not necessarily follow the same pattern. It is also necessary to remember that while these experiments offer a good proxy to quantify risk and measure the efficacy of potential interventions, the dose of airborne viral inoculant in-vivo remains unknown. It should also be remembered that our results so far, apply to the operating theatre under laminar air flow conditions. The apparatus and testing protocol is deployable to all clinical areas, enabling assessments of airflows to be made throughout the hospital in a wide range of clinical scenarios eg ED, ICU, general wards, waiting rooms. Future work will involve the replication of these experiments in the above locations. Further work on engineering controls using high flow local extraction should focus on scalable solutions for other clinical areas.

Our study shows that based on the airflows stated, in real clinical environments, high flow local extraction is a viable solution for removing exhaled aerosols. Further, by qualitative and quantitative laser assessment we have determined the optimal positioning of the high flow local extraction device for maximum efficacy. As the plume could be seen dispersing throughout the whole grid area assessed, although fleeting and of low cumulative probability, it was felt that no zone could be nominated as safe in the absence of HFLE. These conclusions were validated by particle counts at multiple high risk locations within the airflow. We have demonstrated this using a simple, easily engineered high flow local extraction device that utilises existing theatre ventilation and filtration. We believe such a device is easily reproducible and scalable worldwide throughout hospitals.

## Supporting information

Video of standard laminar flow room under laser light demonstrating the plume in multiple axes. 1:54

Video of room with extractor @ 25cm, comparing to matched standard views 1:35

## Data Availability

Full data sets and raw video files are available on request

## Acknowledgements and Statements

We would like to acknowledge the strong support of our anaesthetic and engineering department at Fiona Stanley Hospital. The University of Western Australian has been an invaluable partner in enabling this study.

Financial Support: None reported

All authors contributed equally to the production of this manuscript.

There are no competing interests declared.

*Potential conflicts of interest*. Author L.Marriott has filed a patent (royalty free) on the concept of in-theatre high flow local extraction. All other authors report no conflicts of interest relevant to this article.

The testing conducted in this research project did not involve any patients or members of the public.

This manuscript is an honest, accurate, and transparent account of the study being reported; no important aspects of the study have been omitted; and any discrepancies from the study as planned have been explained.

All data sets in video form are available https://aerosol.fsfhanaesthesia.com/

## Manuscript Appendix

## Appendix 1

### Videos

● Video of standard laminar flow room under laser light demonstrating the plume in multiple axes. 1:54

● Video of room with extractor @ 25cm, comparing to matched standard views 1:35

### Figures and Illustrations

Supplementary Figure 1: CDC hierarchy of controls for risk mitigation.

Supplementary Figure 2: Laser image pre-data processing, at +15cm from midline in the Sagittal plane, showing plume dispersing cephalad under influence of laminar flow.

Supplementary Figure 3: Room optimal airflow design, and noted risks of shortcutting with poor design.

Supplementary Figure 4: Negative pressure Inspiration Gas Exhaling Lung (NIGEL) simulator Flow Volume loops and waveforms.

Supplementary Figure 5: Particle meter assessment being conducted

Supplementary Figure 6: Location of Particle testing

Supplementary Figure 7: Particle counts throughout test. Locations A-D

Supplementary Figure 8: Schematic for RIVET setup. Repurposing Integrated Ventilation for Extraction in Theatre.

Supplementary Figure 9 : Schematic for RIVET Exhaust vent cover version 2.0. Showing sliding louvre system, and hose attachment

Supplementary Figure 10 : Schematic for RIVET Exhaust vent cover version 2.0. Showing sliding louvre system, and hose attachment RIVET Extractor vent cover in situ, affixed to the wall in an airtight yet temporary fashion using duct tape.

**Supplementary Figure 1:**
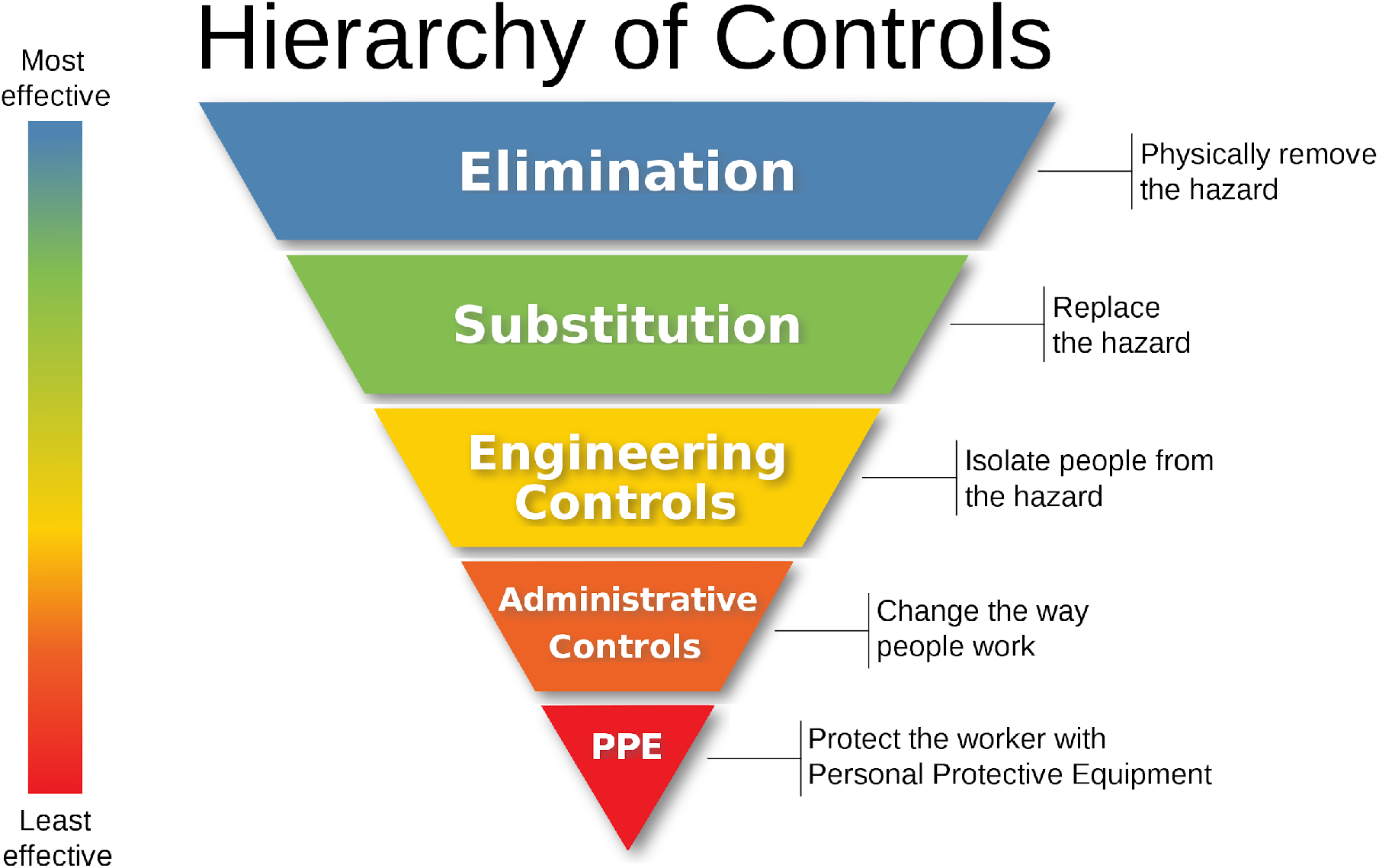
https://www.cdc.gov/niosh/topics/hierarchy/default.html

**Supplementary Figure 2:**
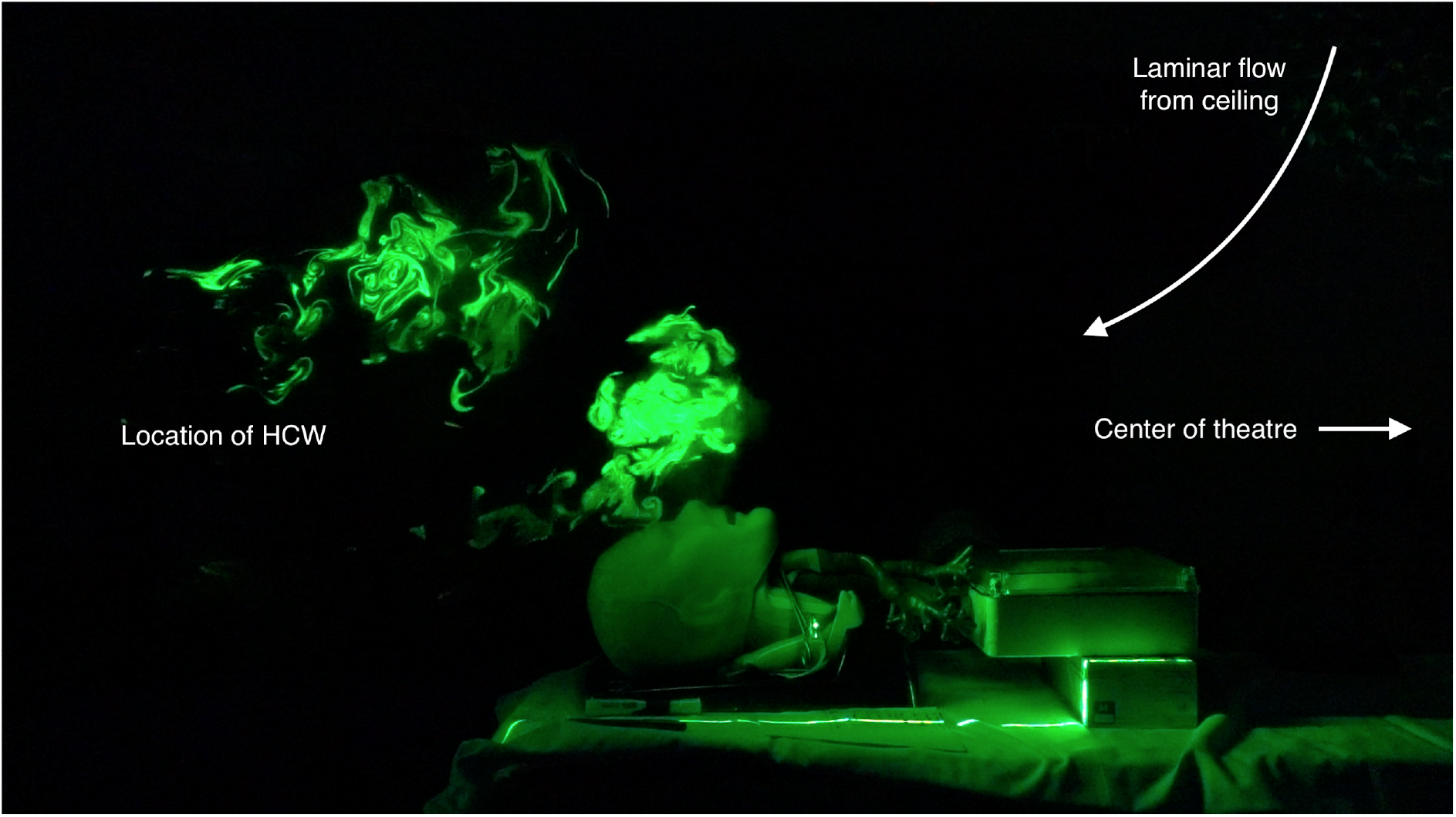
Laser image pre-data processing, at +15cm from midline in the Sagittal plane, showing plume dispersing cephalad under influence of laminar flow.

**Supplementary Figure 3:**
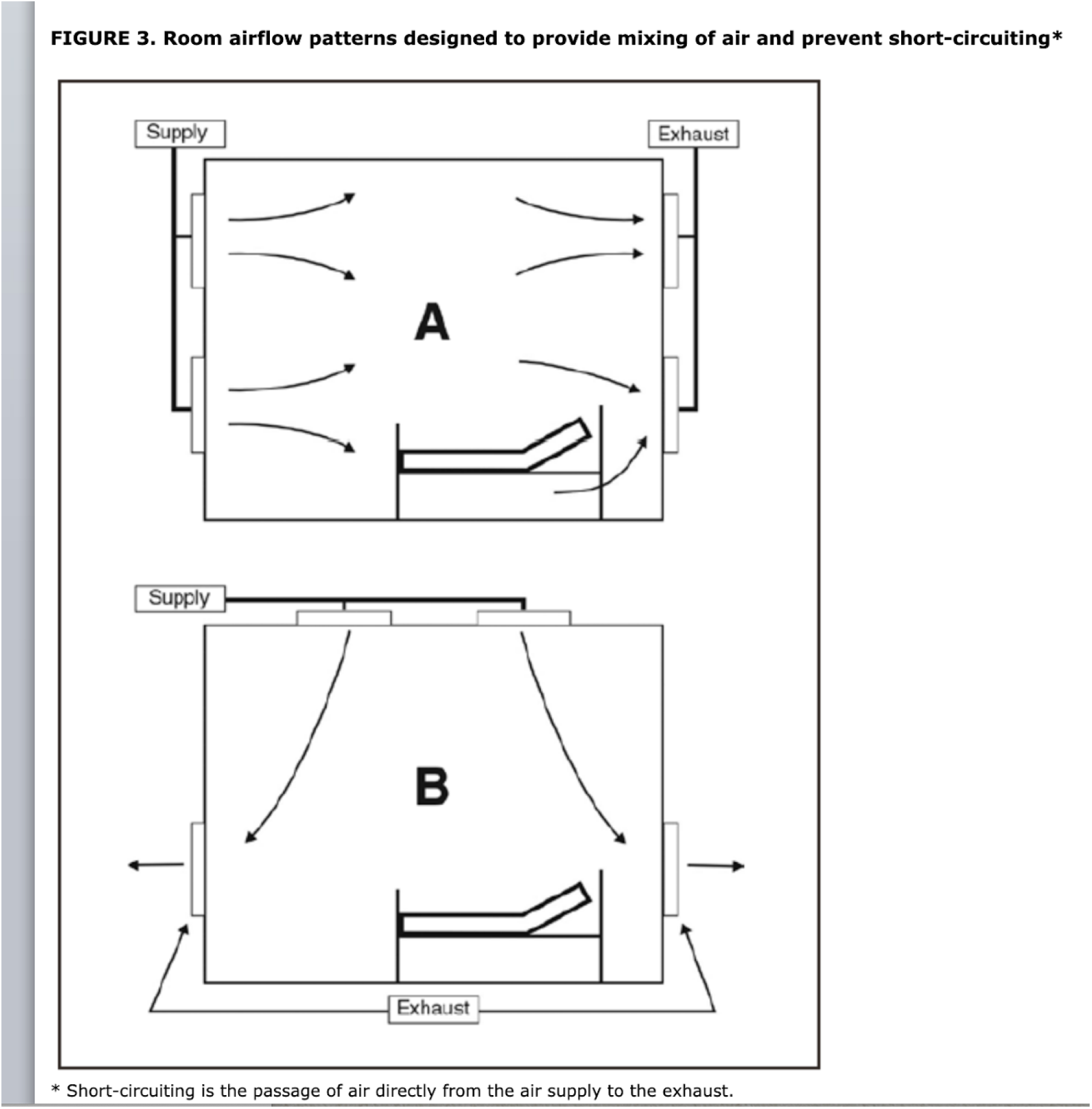
Guidelines for Preventing the Transmission of Mycobacterium tuberculosis in Health-Care Settings, 2005 MMWR 2005;54(No. RR-17): Room optimal airflow design, and noted risks of shortcutting with poor design.

**Supplementary Figure 4:**
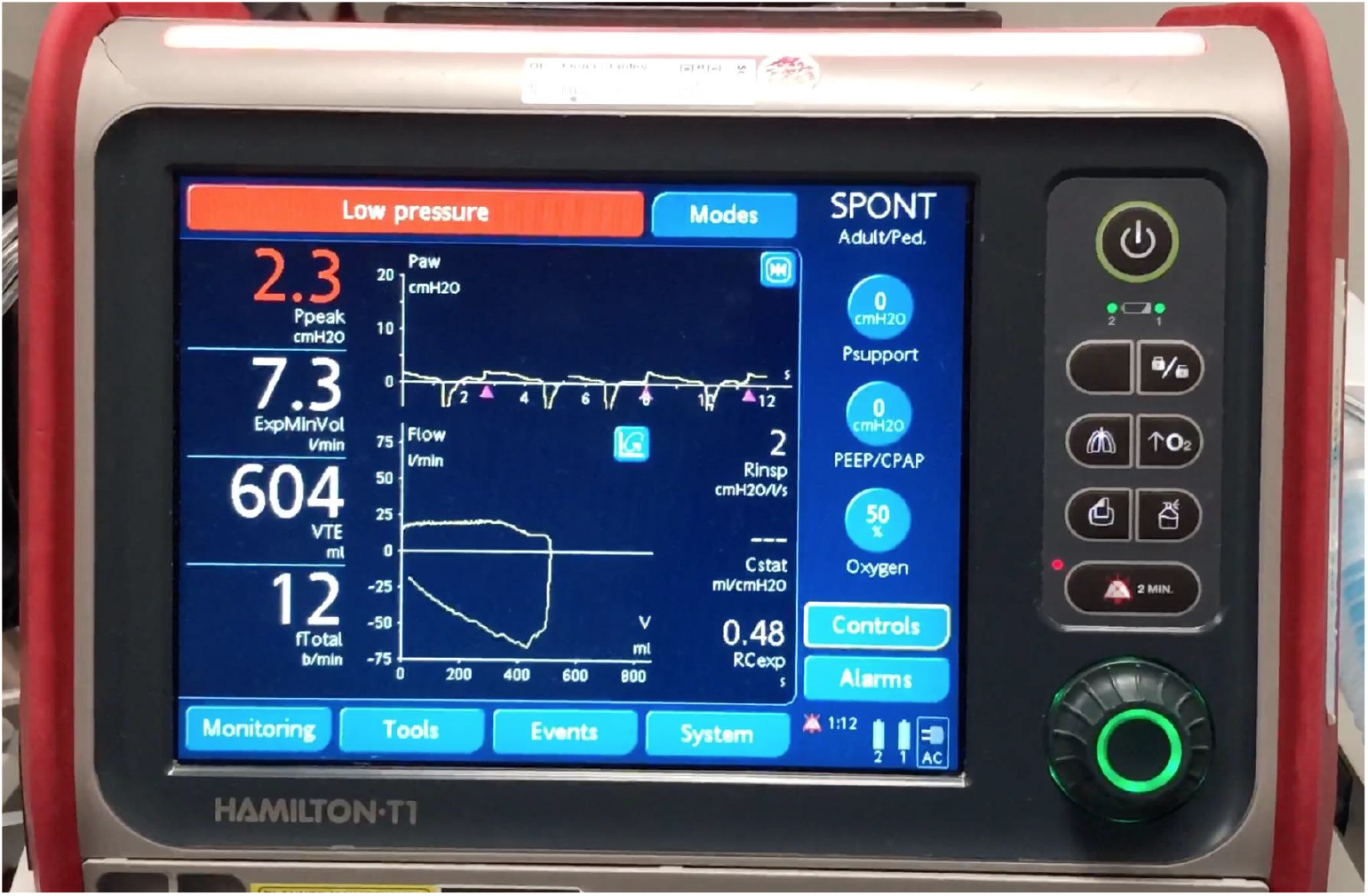
Negative pressure Inspiration Gas Exhaling Lung (NIGEL) simulator Flow Volume loops and waveforms^(b)^ Flow/Volume loop produced by NIGEL set to Vt 600ml, RR 12.

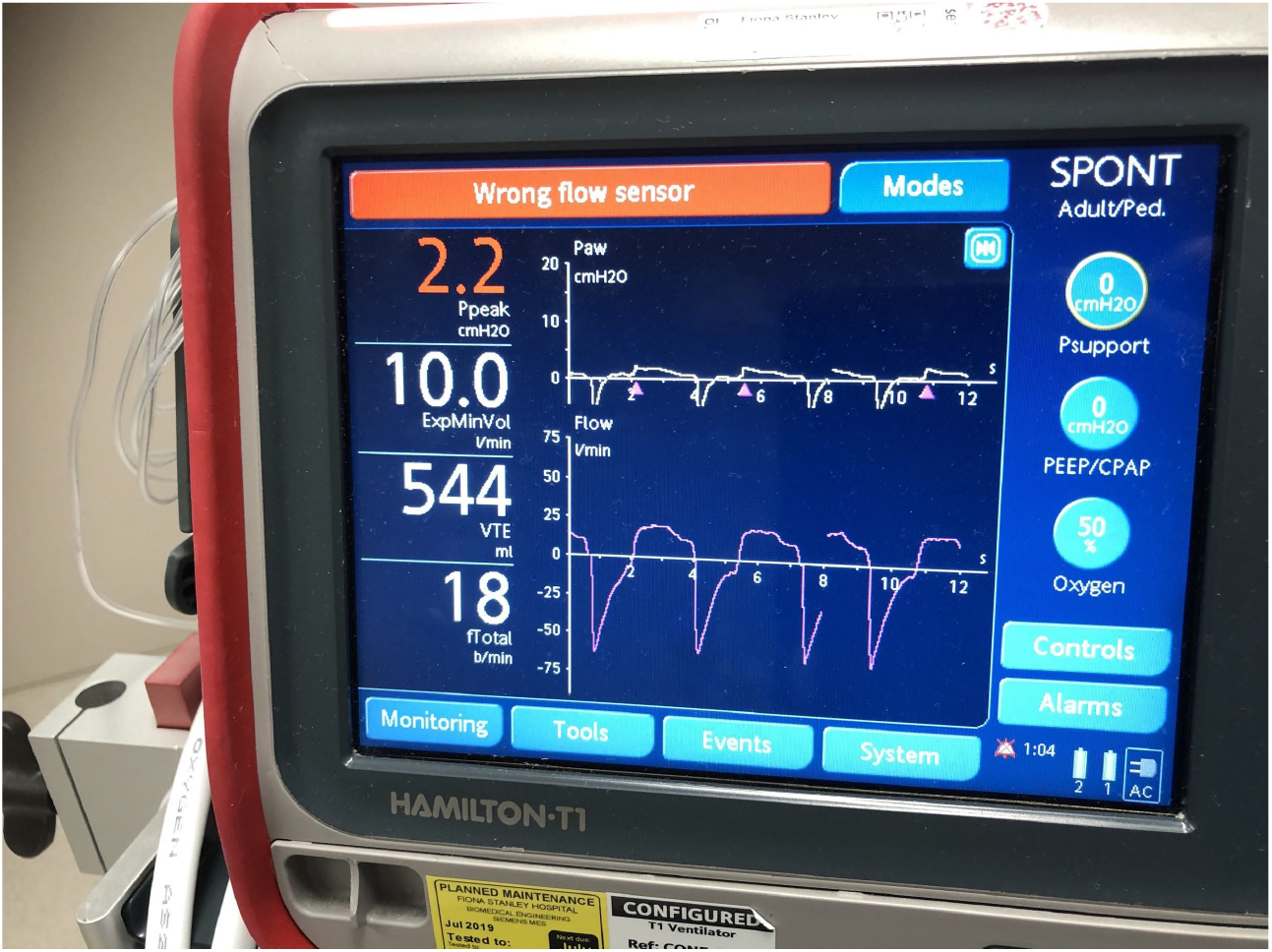
Flow waveform produced by NIGEL set to Vt 650ml, RR 18.

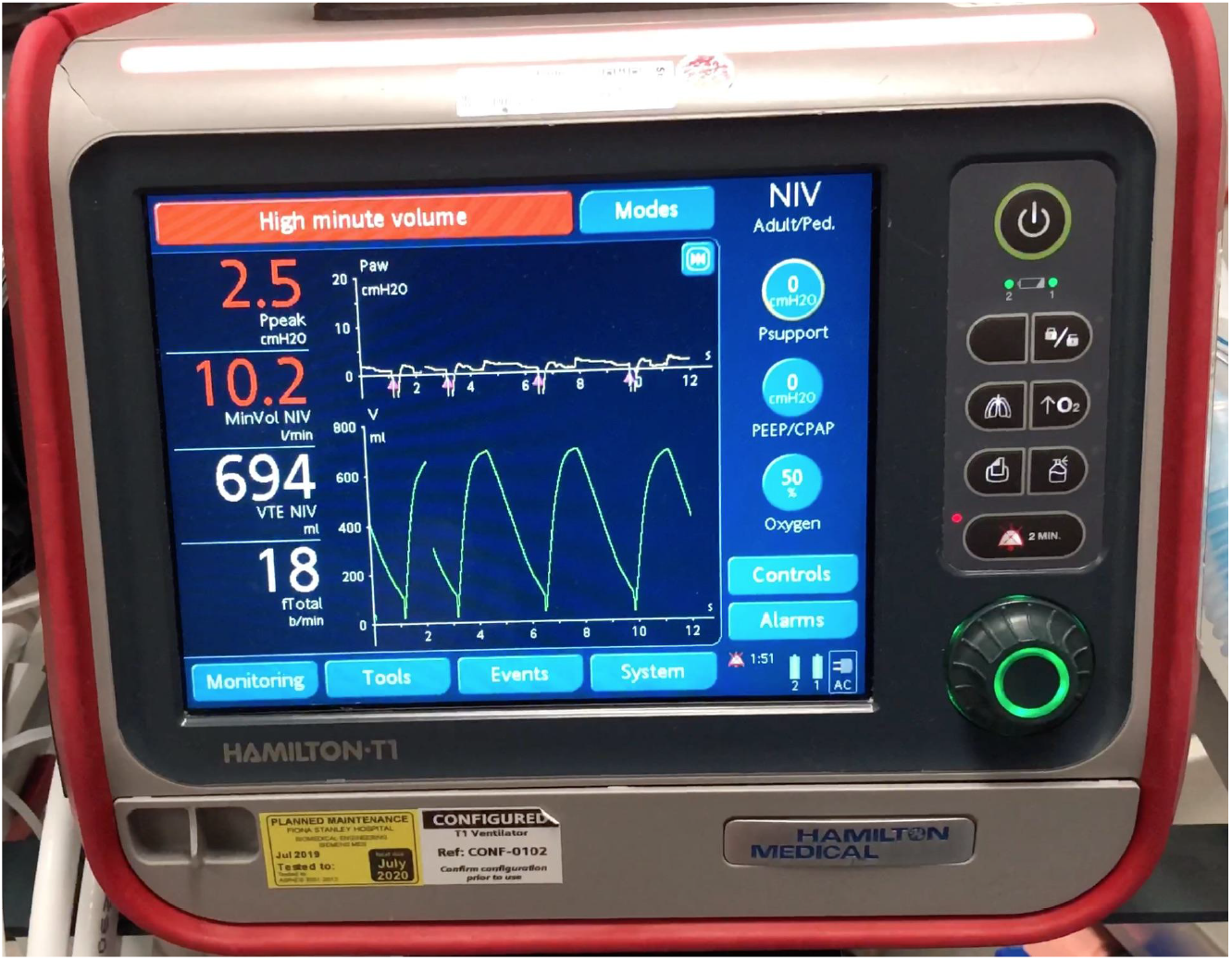
Volume waveform produced by NIGEL set to Vt 700ml, RR 18, during a trial of administering NIV to the lung simulator (current settings: zero support)

**Supplementary Figure 5:**
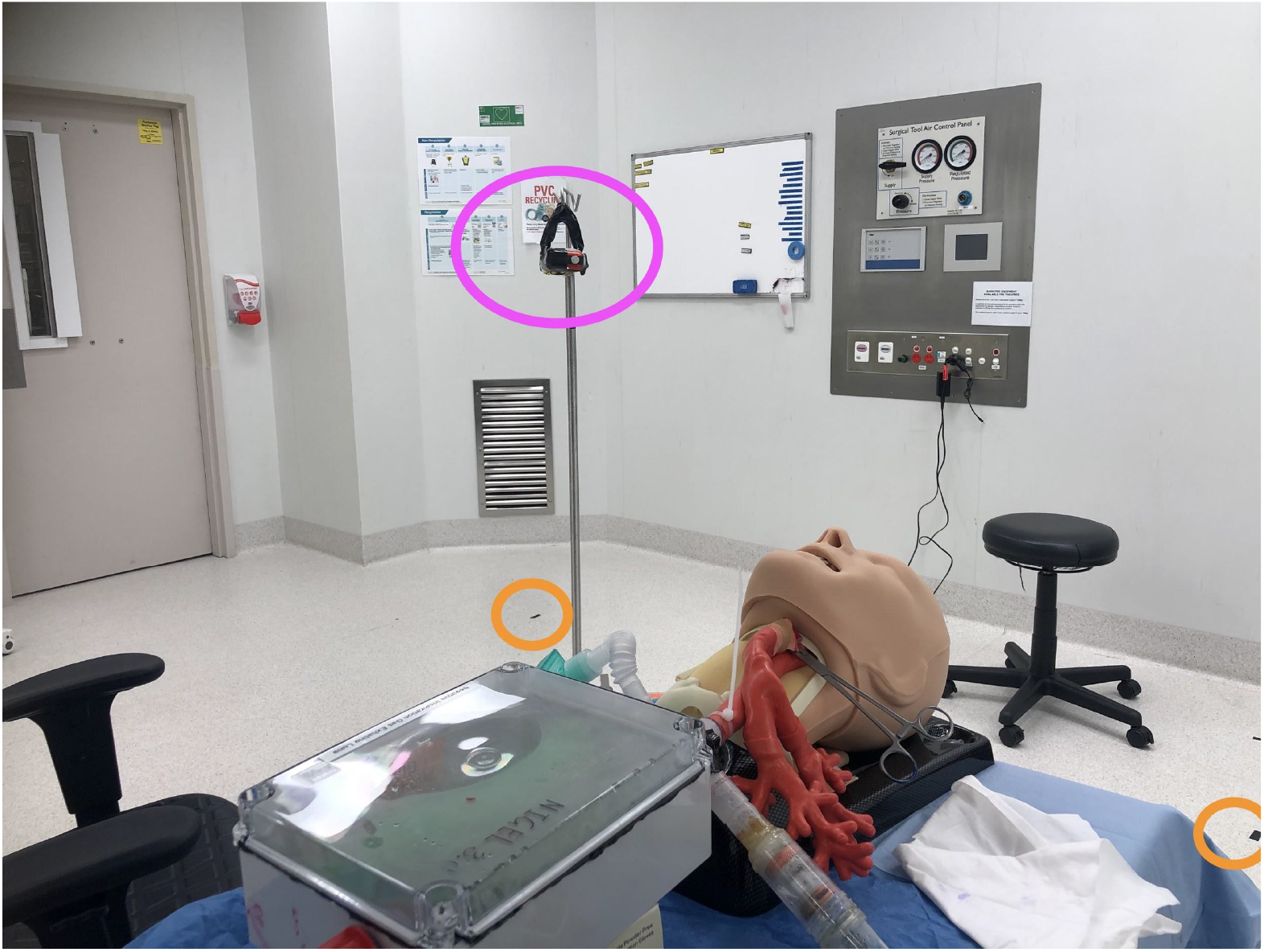
Particle meter assessment being conducted. TESTO DiSCmini particle meter^(g)^ seen here diagonal to the patient, in line with the wall outflow vent, 160cm from the floor (Pink). Other reference locations can been seen in the Sagittal plane of the patient and diagonal (Orange)

**Supplementary Figure 6:**
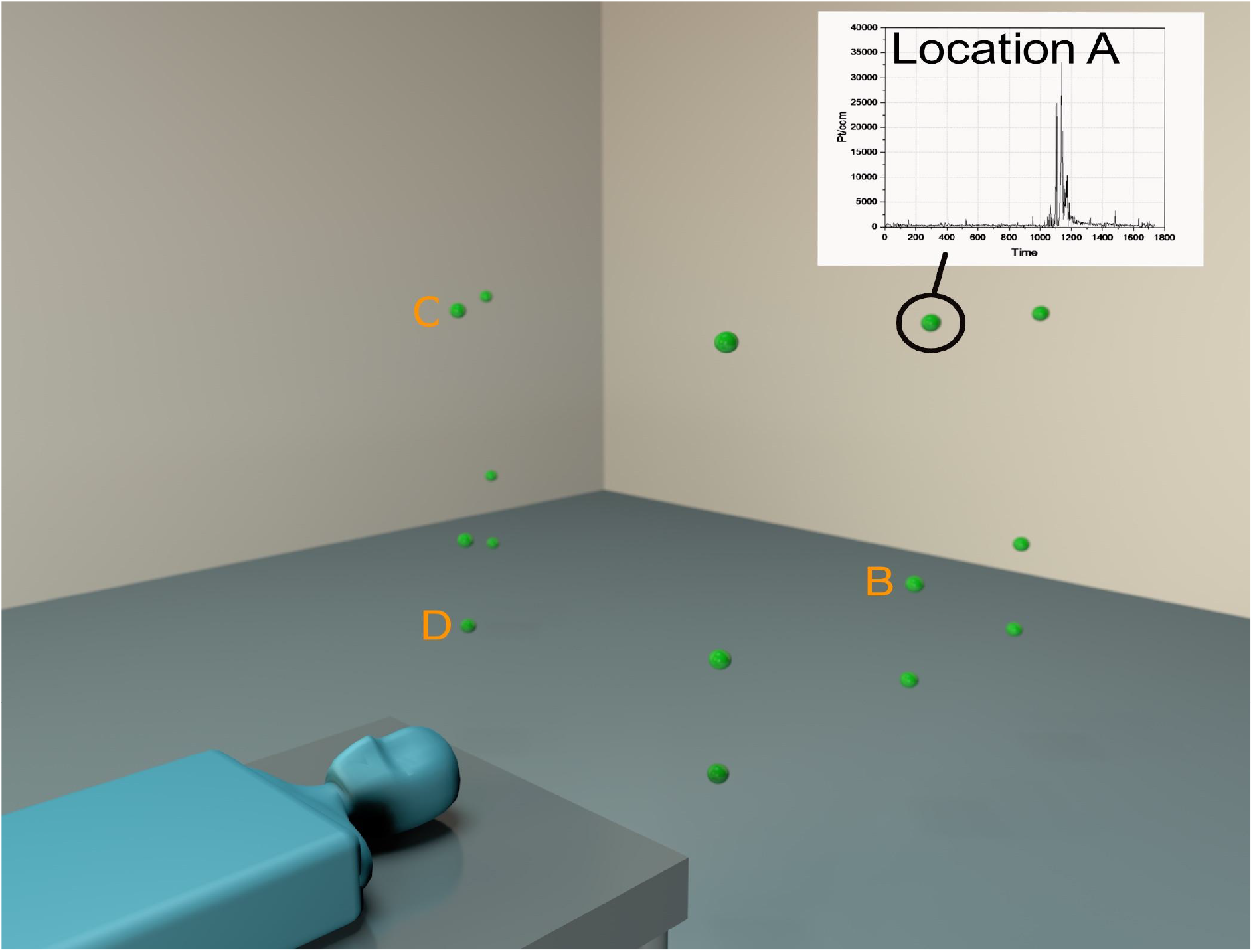
Location of Particle testing Location A: Height of 160cm, Inline, @ 170cm from the patient Location B: Height of 80cm, Inline, @ 170cm from the patient Location C: Height of 160cm, Diagonal, @ 170cm from the patient Location D: Height of 50cm, Diagonal, @ 170cm from the patient

**Supplementary Figure 7:**
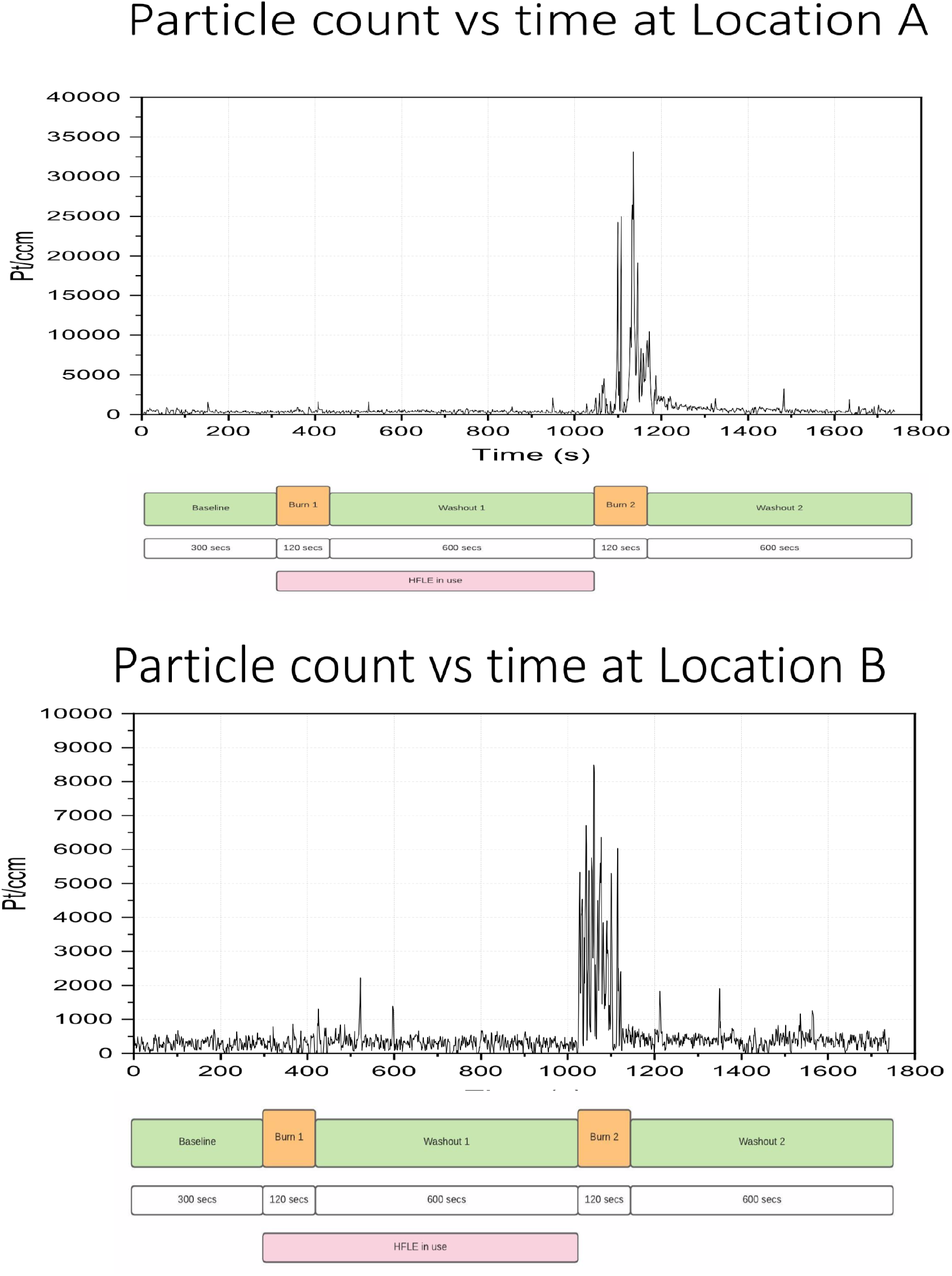
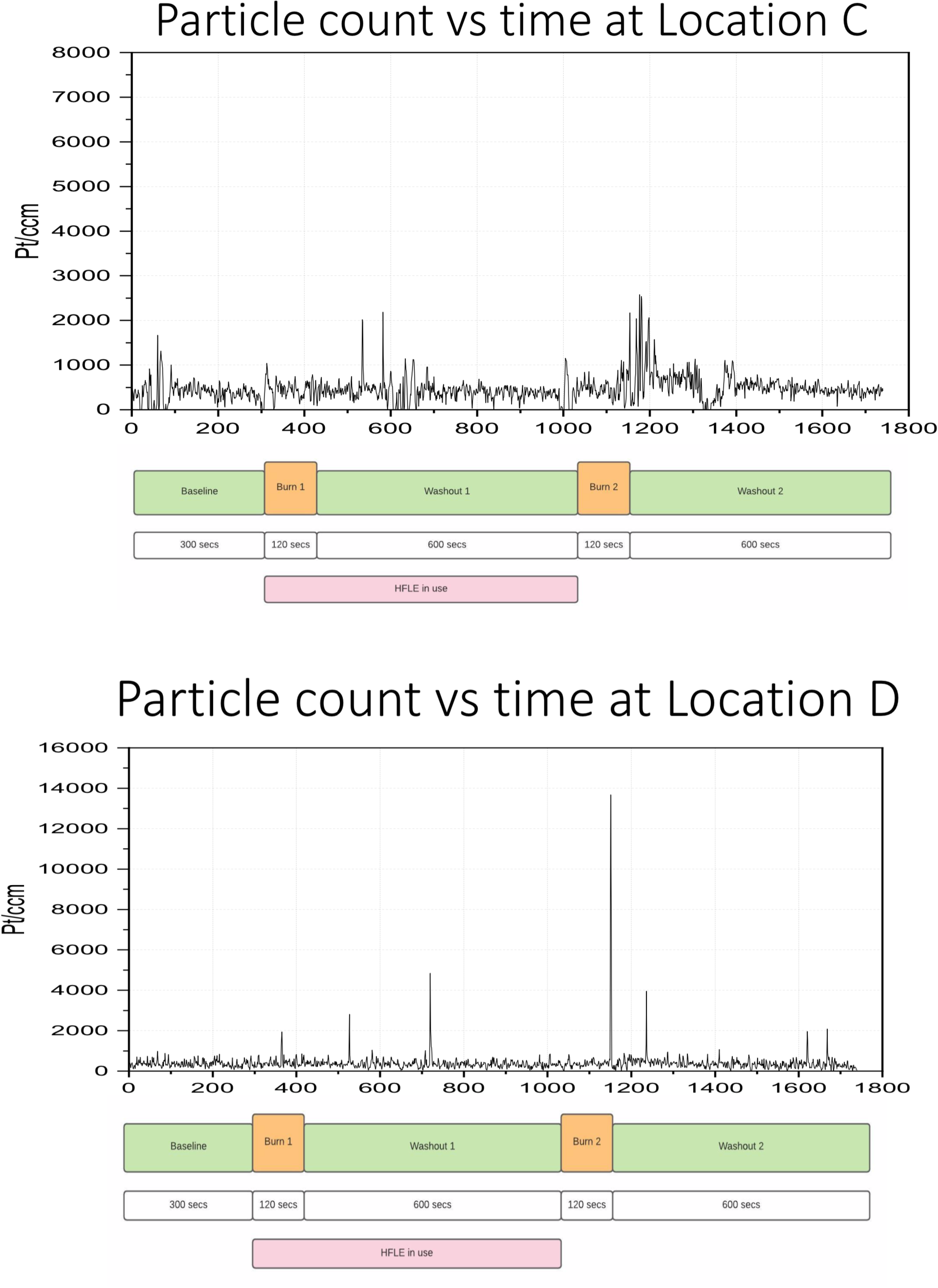
Particle counts throughout the test. Locations A-D

**Supplementary Figure 8:**
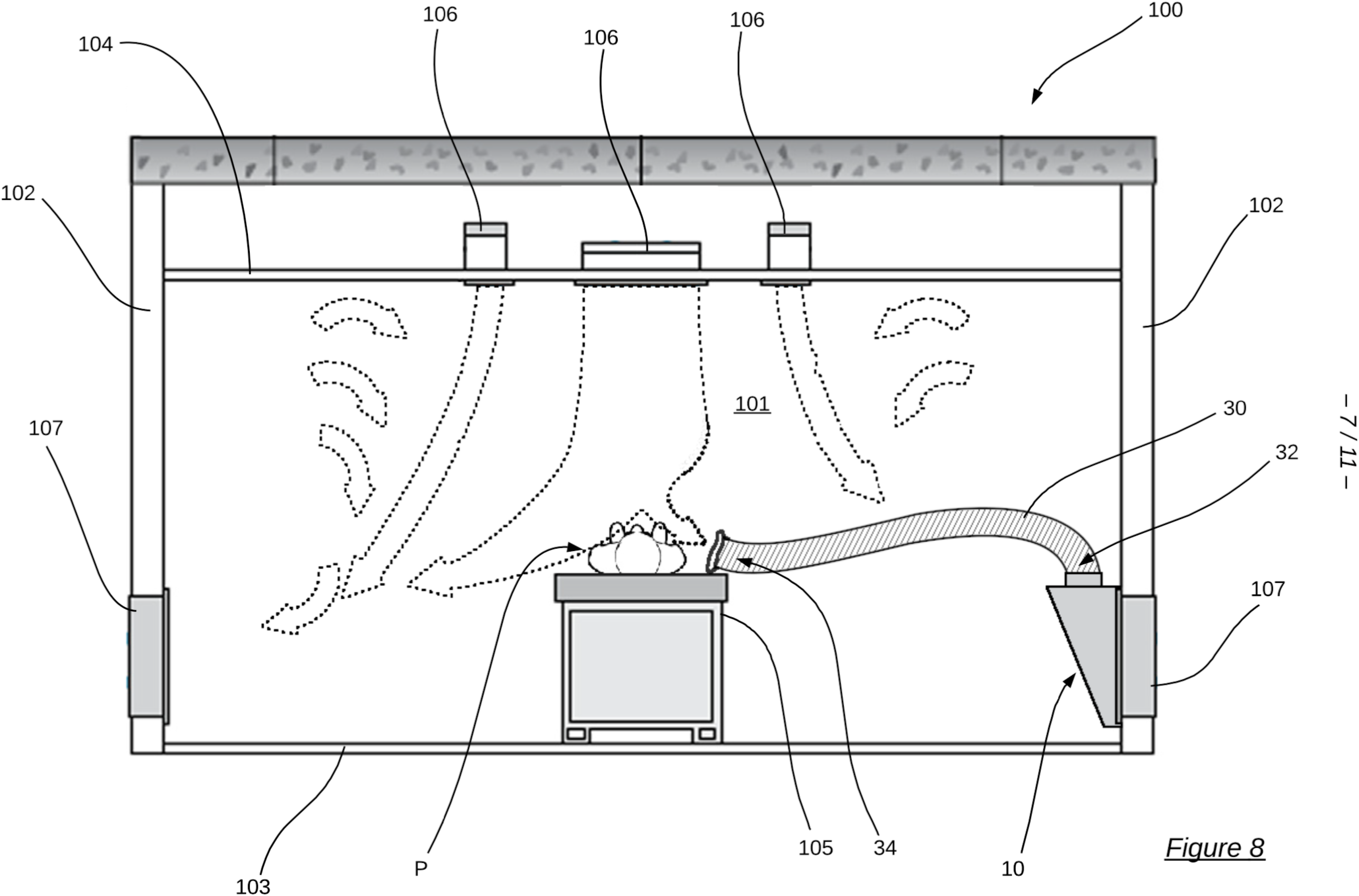
Schematic for RIVET setup. Repurposing Integrated Ventilation for Extraction in Theatre.

**Supplementary Figure 9:**
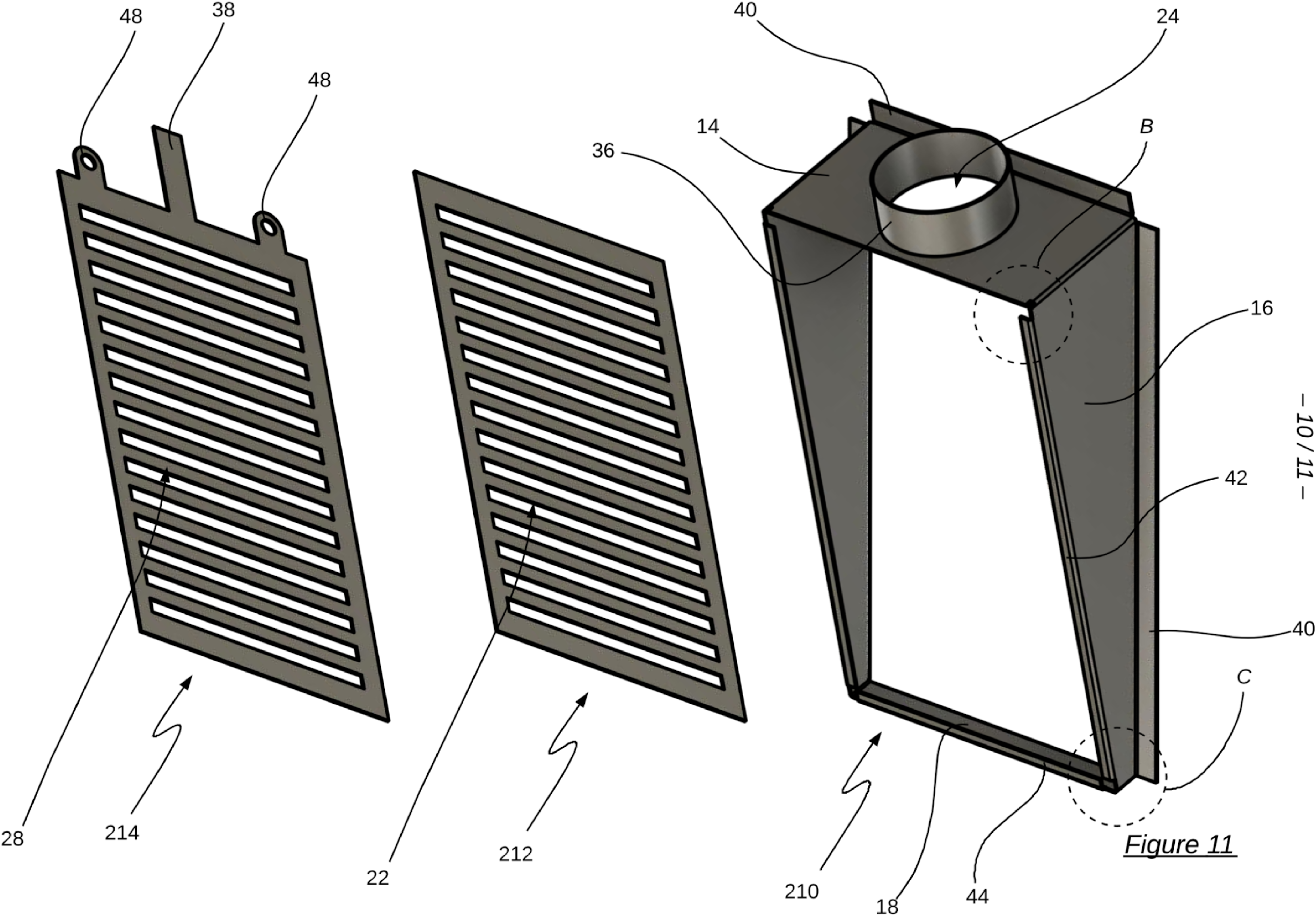
Schematic for RIVET Exhaust vent cover version 2.0. Showing sliding louvre system, and hose attachment

**Supplementary Figure 10:**
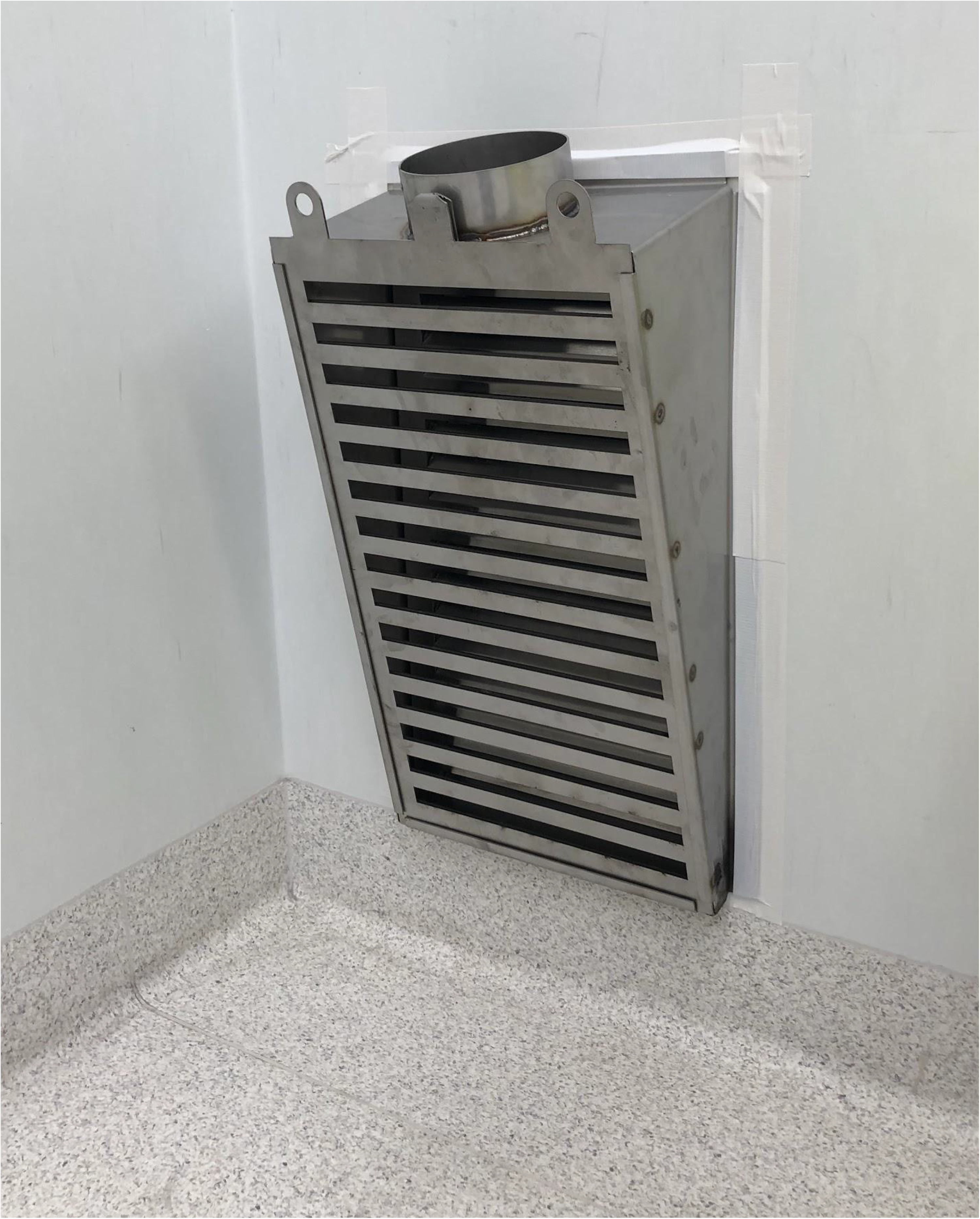
Schematic for RIVET Exhaust vent cover version 2.0. Showing sliding louvre system, and hose attachment RIVET Extractor vent cover in situ, affixed to the wall in an airtight yet temporary fashion using duct tape.

## Appendix 2

### Operating Room physical parameters

Volume 177m3 (8×7.4×3), with 39 air changes per hour (ACH) delivered through eight central ceiling air HEPA diffusers.

Inflow air of 1940l/sec at an average velocity of 0.97 m/s was supplied. This maintained a 12Pa positive pressure relative to the entry corridor, with circulating air velocity at the surgical height reference 0.34 m/s.

Air temperature was maintained at 18°C with 45% relative humidity.

Air was extracted through four floor level vents each equipped with HEPA filters. Prototype high flow local extraction device flow was measured to be 62L/sec at the intake cowling, with velocity 3.5m/sec

### The test apparatus

The Negative pressure Inspiration Gas Exhaling Lung (NIGEL) simulator is made from an airtight transparent 5 liter plastic box drilled to accommodate 3 ports made from standard 19mm Male to Male connectors, sealed in place with structural adhesive at either end of the box.

A primary lung is mounted inside the box (constructed of a 1L paediatric green ventilation bag) connected to the distal port (1). Externally the distal port is connected to a positive pressure ICU ventilator (Mindray). The proximal port (2) is connected externally to an Airway Mannequin head (TruCorp) via the tracheal tubing using sections of paediatric anaesthetic circuit.

The ventilator is programmed to ventilate the secondary lung, with an inverse I:E ratio due to the reciprocal nature of the NIGEL. Inflation of the primary lung displaces the remaining gas volume out of the box via the proximal port, and on deflation the box ‘inspires’. Altered inflation times, ramp, and a maximal driving pressure approaching 100cmH2O is used on the primary lung.

Pressure volume loops were recorded using a second anaesthetic machine flow sensor set to spontaneous mode on the proximal port. See Supplementary Figure 4.

The TruCorp AirSim Advance Bronchi X intubation and bronchoscopy training head^(a)^ has anatomically realistic naso & oropharyngeal contours, allowing correct airflow velocities due to volumes exhaled being determined by outflow tract diameters and structures such as the tongue, turbinates and soft palate.

100% Propylene Glycol is vaporised using an E-cigarette coil device^(c, d)^ which is mounted via a T piece onto the mannequin trachea. The 0.2 Ohm E-cigarette coil is powered by a modulation box delivering a constant wattage (120W nominally for our experiments), at 4.7-5.2V. Vaporisation occurs in the inner chamber, saturating the airspace with Propylene Glycol. This is placed on a T-piece connected to the test apparatus distal airway. Aerosol is entrained into the test apparatus during inspiration. Supplemental airflow delivered by an aquarium pump^(e)^ at 2.4lt/min provides the air supply to be saturated, thereby preventing coil burnout.

### Lazer imagery

The field of view was illuminated by a 5-Watt continuous wave Argon-ion laser, with a scattering angle of 38 degrees. The flow field was recorded by a digital camera (Sony Cyber-shot RX100 V^(f)^), with a resolution of 1920 x 1080 pixels and frame rate of 50fps. Both the laser and the camera were mounted on tripods. The camera was positioned perpendicular to the axis of the laser beam.

The individual frames were elicited and converted into grey-scale images for probability analysis. Those processes were performed by Matlab code which was designed explicitly for this study. Reference images must be taken before each test to eliminate any permanent background lighting. By subtracting the reference image, each frame represented the intensity of the light sheet corresponding to the smoke concentration. The images were normalized with the highest intensity value in the smoke, and the results were applied to establish the particle concentration topologies.

A similar technique was described previously by D Hui

Laser imagery tests were first conducted without high flow local extraction in operation. The high flow local extraction device was then positioned on the mannequin’s “chest” at 20cm from the mouth, and then at three increments of five cm caudally to a maximum of 35 cm from the mouth. In each of these positions tests were completed in three sagittal slices and five axial slices. A total of 52 laser and device configurations were tested across both axes and four extractor positions to most effectively map plume behaviour both with and without high flow local extraction.

### Particle meter

TESTO DiSCmini^(g)^ records the particle number in a time resolution of 1 second, in the range from 10 to 700 nm, and the mean particle size in the range from 10 to 300 nm

Particle size: 10 to 700 nm (absolute. for particle number)

10 to 300 nm (Modal. for mean particle size)

Particle concentration: 1000 to 1000000 Particle/cm^3^ (typical values)

Flow rate: 1.0 L/min ± 0.1 L/min (16ml/sec)

Units; particles/cm^3^

Mounted at 5 fixed points throughout the room.

These locations were determined by direct observation of propylene glycol aerosol behaviour in theatre airflows, coinciding with the likely locations of staff positioned downstream of the airflow.

1. directly inline with the sagittal axis of the test apparatus, at distances of 90, 170, and 230cm from the mannequins mouth.
2. diagonally toward the corner exhaust vents at distances of 160 and 270cm.

In each of these locations measurements were made at three heights of 160cm, 80cm and 50cm from the floor.

Prior to each test, all room doors were opened fully and the room was left empty for 20 minutes to allow clearance of residual aerosols from previous tests. Room doors were then closed and secured to prevent aberrant/inconsistent airflow during testing, and the research team remained stationary within the room.

## Appendix 3

### Prototype HLFE device

Fiona Stanley Hospital theatres airflow system is of industry standard, having a centrally mounted ceiling inflow, and 4 floor level corner outflows with both of these being fan forced. The balance of these is maintained to create a pressure above the ambient pressure of surrounding areas by approximately 10 pascals resulting in a positive pressure theatre to prevent airborne contaminants entering the surgical field. Flow rates result in over 20 air changes per hour, of a typical 150m3 room, with a quarter of this being pulled from each corner duct.

Downstream is HEPA filtration before exiting the building system.

The prototype Extractor works by diverting normal room extraction from one such corner duct, in a process we have termed RIVET (Repurposing Integrated Ventilation for Extraction in Theatre). One corner vent is shrouded with a custom built, baffled plenum box, with an intake nozzle on the top surface. It is attached to the wall with duct tape and is easily removed and cleaned. The box is constructed of stainless steel and the front has louvres which can be closed to engage the extraction from the intake nozzle or opened, to allow “normal” room extraction. Simple air duct tubing (15cm diameter, 5m length) is attached to the intake nozzle and brought across the room with the “patient” end positioned on the chest close to the airway. A cardboard flange is attached at this end to form a funnel shaped cowling. The stainless steel box may be easily cleaned, however the hosing and nozzle are disposed of after use.

Supplementary Figure 9: Schematic for RIVET Exhaust vent cover version 2.0. Showing sliding louvre system, and hose attachment

The prototype HFLE device for use in theatre can be turned on (baffles closed) for localised extraction from the airway during AGPs, and then turned off (baffles open) to restore normal room “Laminar flow” if required, for the duration of surgery. It is hypothesised that normal airflow patterns of laminar flow over the surgical field are only needed during the sterile surgical time period of the patients theatre journey, and that at all other times the airflow could be contained to have a localised extraction near the patients face, particularly during intubation and extubation or throughout non sterile procedures such as bronchoscopy/gastroscopy.

This could provide a greater reduction of room contamination.

Supplementary Figure 10: Schematic for RIVET Exhaust vent cover version 2.0. Showing sliding louvre system, and hose attachment RIVET Extractor vent cover in situ, affixed to the wall in an airtight yet temporary fashion using duct tape.

During our testing the prototype high flow local extraction device flow was measured to be 62L/sec at the intake cowling, with velocity 3.5m/sec

*Other devices, as yet unvalidated and or requiring extensive building modification:*

https://www.nederman.com/en-us/applications/aerosols-suction

https://about.unimelb.edu.au/newsroom/news/2020/april/researchers-design-ventilation-hoods-for-hospital-beds-to-help-contain-covid-19-spread

https://vapotherm.com/clinical/felix-1-white-paper-computational-fluid-dynamic-modelling-of-particle-capture-possible-use-of-a-modified-face-tent-as-a-facial-scavenger/

## Footnotes

a. TruCorp AirSim Advance Bronchi X, TruCorp Ltd, Craigavon United Kingdom

b. Hamilton T1, Hamilton Medical, Bonaduz Switzerland

c. FreeMax Mesh Pro, FreeMaxVape Shenzhen China

d. Vaporesso Gen Nano, Shenzhen Smoore Technology Limited

e. Fish Pump, Aqua One Precision Air Pump 2500 150L

f. Sony Cyber-shot RX100 V, Sony Corporation Tokyo Japan

g. TESTO DiSCmini, Testo Australia Ltd, Melbourne Australia

